# Reduced loss aversion in subclinical negative symptoms and hypomania

**DOI:** 10.1101/2020.01.27.20018119

**Authors:** Federica Klaus, Justin Chumbley, Erich Seifritz, Stefan Kaiser, Matthias Hartmann-Riemer

## Abstract

Loss aversion is a behavioral phenomenon that describes a higher sensitivity to losses than to gains and influences decisions. Decision-making is altered in several psychopathologic states, such as in the two symptom dimensions of hypomania and negative symptoms. It has been argued that progress in our understanding of psychopathology requires a reorientation from the traditional, syndrome-based perspective to a more detailed study of individual constituent symptoms. In the present study, we made careful efforts to dissociate the relationship of loss aversion to negative symptoms, from its relationship with hypomanic symptoms. We selected a sample of 45 subjects from a healthy student population (n = 835) according to psychopathologic scales for hypomania and negative symptoms and stratified them into a control group (n = 15), a subclinical hypomania group (n = 15) and a negative symptoms group (n = 15). Participants completed a loss aversion task consisting of forced binary choices between a monetary gamble and a riskless choice with no gain or loss. We found, that these two symptom dimensions of hypomania and negative symptoms have a similar inverse relation to loss aversion as demonstrated by analysis of variance. Further research is warranted to describe the underlying psychological and neurobiological mechanisms at play. Given the partially opposing nature of hypomania and negative symptoms it further needs to be elucidated whether they are linked to loss aversion via dissociable mechanisms.

## Introduction

Negative symptoms (apathy, diminished expression) and (hypo-) mania are two severe and to some extent opposing symptom dimensions that occur in several psychiatric diseases, such as schizophrenia, depression or bipolar disorder [1]. Both symptom dimensions significantly impair everyday functioning [2,3]. Moreover, they have been linked to altered decision-making behavior [4–7], which might partially cause and/or maintain observed symptoms.

Behavioral economics provides powerful methods to investigate variance in decision-making. Recently, these methods have been fruitfully applied in the study of psychopathology [8–10]. Negative symptoms and (hypo-) mania are typically associated with a reduction or an increase, respectively, in the frequency and vigor of goal-directed behavior. Valuation of losses and gains and their associated uncertainty constitute core decision-making processes guiding adaptive goal-directed behavior. Thus these processes might be especially relevant regarding negative and (hypo-) manic symptoms, since clinical observations show an increase in goal-directed behavior and associated investment of effort in mania [11,12] and a decrease in goal-directed behavior and invested effort or motivation in negative symptoms [13,14], both resulting in an impairment of effort-cost computations.

A well-replicated phenomenon in behavioral economics is *loss aversion*, which describes a higher sensitivity to losses than to gains, observable in the general population. In other words, the value of an object is judged higher when it is lost compared to when it is gained, depending on the reference point [15,16]. Loss aversion may be understood from an evolutionary point of view [17]: when one’s survival is at risk, marginal losses prove more critical for reproductive success than marginal gains [18] and in an environment with low resources, humans are cognitively biased to ensure that they do not fall below some minimal threshold of resources necessary for survival [19]. A marked miscalibration of an individual’s loss aversion might impede their social and economic effectiveness and be accompanied by psychopathology.

A reduced or absent sensitivity to loss in schizophrenia patients has been reported [20–22]. Moreover, one study found a significant negative correlation between total symptom severity and loss aversion [21]. This is opposed to the concept that an increased loss aversion could contribute to a reduced goal-directed behavior, which in turn manifests itself as negative symptoms. However, no study so far has addressed potential links to specific symptom dimensions, such as negative symptoms. Moreover, to our knowledge no study has investigated loss aversion in (hypo-) manic states, which is surprising, considering that the diagnostic criteria for manic episodes [1] include impulsivity and disregard for the potential losses and risks accompanying one’s actions. A study in euthymic bipolar patients observed a significant decrease in loss aversion in a computerized decision-making task in the patient group compared to healthy controls [23]. Another study found no performance deficit in manic bipolar patients during a probability-based gambling paradigm dependent upon risk-taking situations aimed at maximizing gain and minimizing loss [24]. However, the direct link between (hypo-) manic or negative symptom dimensions and the degree of loss aversion remains to be elucidated.

Importantly, features of negative symptoms and (hypo-) mania can be found not only in severely ill patients, but vary along a continuum in the non-clinical population [25,26]. These dimensional approaches to psychopathology hypothesize that clinical and sub-clinical symptom expression should at least in part be associated with similar underlying mechanisms. Thus, the investigation of the extreme individuals within the “normal” population, being more accessible to study, offers a powerful way to dissect sub-clinical and clinical disease mechanisms [27,28].

Here, we aimed to investigate whether elevated negative and hypomanic symptoms in a stratified non-clinical sample are associated with changes in loss aversion using a binary choice task with monetary incentives. Based on the literature and listed symptoms in current diagnostic systems [1,29], we hypothesized that negative symptoms and hypomania would both be associated with a reduction in loss aversion: As summarized above, a reduced or absent sensitivity to loss has been reported in schizophrenia, of which negative symptoms are one of the main psychopathologic constituents. Regarding (hypo-) mania, its clinical definition includes impulsivity and disregard for the potential losses, which is a behavior congruent with a reduced aversion to losses.

## Methods

### Ethics statement

All participants provided informed consent via online assessment. The study complied with the Declaration of Helsinki and the local ethics committee of the Canton of Zurich approved the procedure.

### Participants and procedure

835 students of the University of Zurich (607 male; age: *M* = 24.3, *SD* = 5.49) were recruited through university mailing lists and social media and filled out online trait questionnaires assessing hypomania using the Hypomania Symptom Scale (HPS-30: *M* = 11.1, *SD* = 4.88) [30] and negative symptoms using the negative symptom items of the Community Assessment of Psychic Experiences (CAPE, *M* = 1.62, *SD* = 0.49) [31]. As reimbursement they took part in a lottery for Amazon gift cards (300 Swiss Francs, i.e. ∼300 USD). Exclusion criteria were current treatment for a psychiatric disorder (therapy and/or medication), inclusion criteria was an age between 18 and 55 years. On the basis of this large reference population we defined high negative symptoms / low hypomania (“negative symptom only group”), high hypomania / low negative symptoms (“hypomania only group”), and low negative symptoms / low hypomania (“control group”) target subpopulations. The cut-off for “low” or “high” scores was defined as scores that were below the 40th and above the 60th percentile of the reference population respectively.

In a second step, participants meeting target criteria (total *n* = 45) were invited to the laboratory: 15 participants (60% male; age: *M* = 23.8, *SD* =3.84 years) with low scores on the negative symptom subscale and low scores on the hypomanic personality scale (CAPE score: *M* = 1.11, *SD* = 0.15; HPS-30 score: *M* = 5.73, *SD* =1.71) (control group), 15 participants (73.3% male; age: *M* = 25.9, *SD* = 5.26 years) with high scores on the negative symptom subscale and low scores on the hypomanic personality scale (CAPE score: *M* = 2.23, *SD* = 0.43,; HPS-30 score: *M* = 5.87, *SD* = 1.64) (negative symptom only group) and 15 participants (60% male; age: *M* = 21.8, *SD* = 2.24 years) with low scores on the negative symptom subscale and high scores in the hypomanic personality scale (CAPE score: *M* = 1.15, *SD* = 0.17; HPS-30 score: *M* = 16.93 *SD* = 2.49) (hypomania only group) took part in the decision-making experiment.

The reference sample (n = 835), the stratification, and the recruited study participants are depicted in Fig 1.

**Fig 1.**
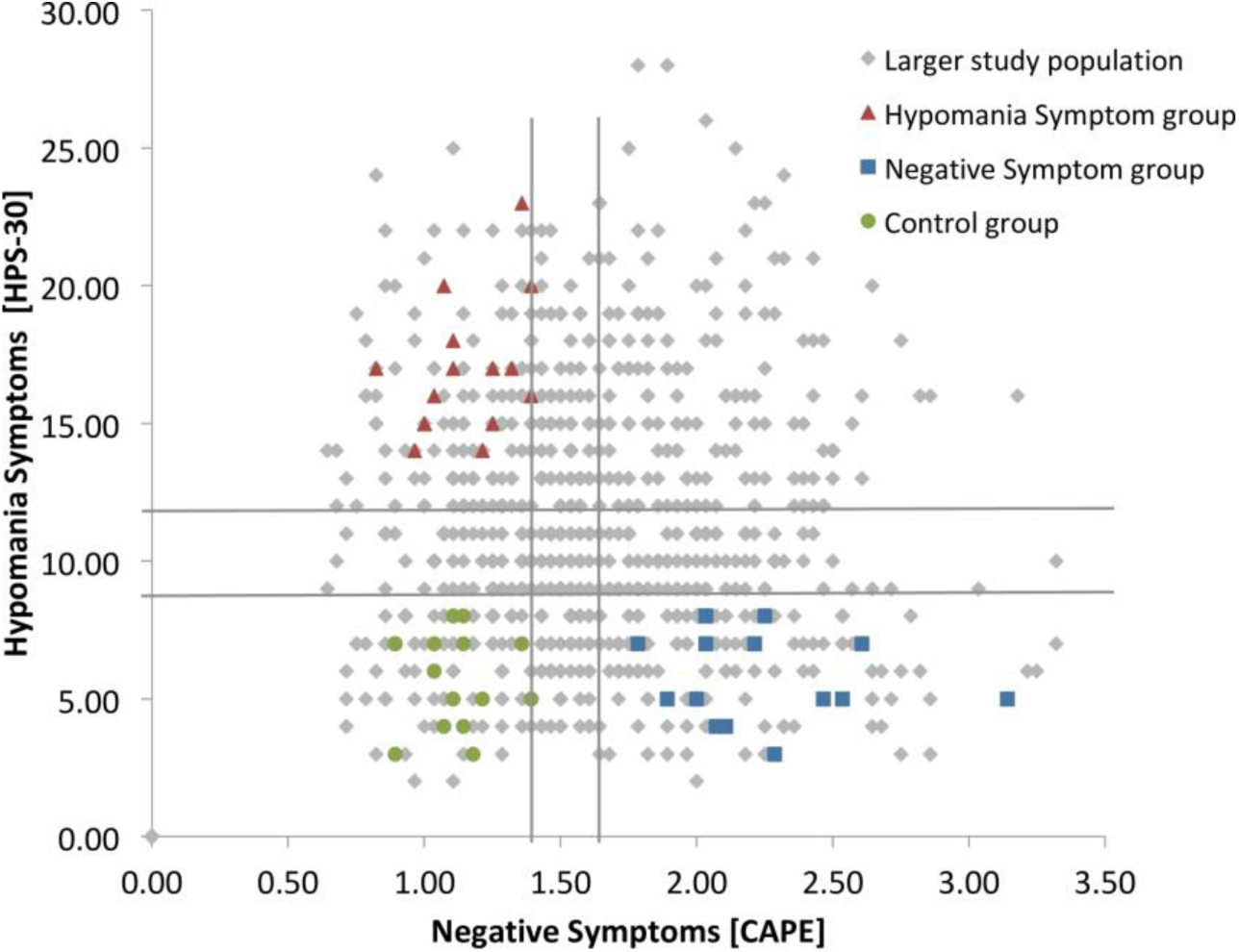
Sample stratification procedure. On the basis of a large reference population (*N* = 835) we defined high negative symptoms / low hypomania (“negative symptom group”), high hypomania / low negative symptoms (“hypomania group”), and low negative symptoms / low hypomania (“control group”) target subpopulations. The cut-off for “low” or “high” scores was defined as scores that were below the 40th and above the 60th percentile of the reference population respectively (grey lines).

### Experimental task

Participants were endowed with 30 CHF at the beginning of the study. Participants then completed a loss aversion task [32], consisting of 20 forced binary choices between a monetary gamble (P=0.5) and a riskless choice with no gain/loss (P=1). Participants were presented on a computer screen three numbers representing amounts of money, two of them representing the possible gain or loss in case the gamble was accepted and one representing 0 CHF in case of a rejection of the gamble. Following standard practice in behavioral economics, subjects knew in advance that their final payment would equal their 30 CHF endowment, plus or minus one of their 20 outcomes, selected at random. The outcome of each chosen gamble was revealed to the subject immediately. At the end of the experiment, they were then paid according to the policy just detailed above, debriefed and dismissed. We used their choices to quantify a subject-specific loss parameter, called λ below. In addition, choices were used to quantify attitudes toward chance (risk aversion ρ) and consistency over choices (logit sensitivity μ).

### Data analysis

We used a three parameter model to estimate choice behavior: Gains were estimated by exploiting equation 1: *u*(x^+^)=x^ρ^, and losses via equation 2: *u*(x^-^)= -λ*(-x)^ρ^. These combine in equation 3: p(gamble acceptance)=(1+exp{-μ(*u*(gamble)-*u*(guaranteed))})^-1^, where *u*(gamble) is the difference between equation 1 and 2. *u* indicates the anticipated utility or “desirability”, x indicates the (monetary) value; ρ is the curvature of the utility function and indicates the risk aversion, λ is the loss aversion coefficient and refers to the multiplicative valuation of losses relative to gains (with λ>1: loss aversion, λ<1: gain seeking, λ=0: gain-loss neutral), p indicates the probability of an event and μ indicates the sensitivity of the participant’s choices to changes in the difference between subjective values of the gamble and the guaranteed fixed amount (consistency). The subject-specific parameter λ, our main outcome representing loss aversion, was estimated from subject-specific expected marginal posteriors. Parameters μ and ρ were estimated in the same way. For details see [32–34]. To determine the between-subject differences, a one-way multivariate analysis of variance using IBM SPSS 25 was performed with λ, μ and ρ as dependent measures and group as independent measure. Age was introduced as covariate, post-hoc testing was performed using LSD (least significant difference).

## Results

### Sample characteristics

The three groups did not differ significantly in gender (*F*(2,42) = .368, *p* = .694), income (*F*(2,42) = .883, *p* = .421), education (*F*(2,42) = 1.451, *p* = .246) or money spent per month (*F*(2,42) = 1.037, *p* = .363). However, a significant group difference regarding age was observed (*F*(2,42) = 4.055, *p* = .025). The hypomania group (age: *M* = 21.80, *SD* = 2.24) was significantly younger than the negative symptom (age: *M* = 25.93, *SD* = 5.26) group with a mean difference of 4.13 years (*p* = .007), while the negative symptom and the hypomania group did not differ in age in comparison to the control group (age: *M* = 23.80, *SD* = 3.84)(*p* = .149 and *p* =. 176 respectively). We therefore used age as a covariate in some analyses below.

### Loss aversion is inversely related to negative symptoms and hypomania

Groups did differ in their loss averse choice behavior (*F*(2,42) = 7.919, *p* = .001). Post-hoc testing revealed that participants with higher negative symptoms or hypomania showed less loss averse choice behavior compared to controls (*p* = .0003 and *p* = .025 respectively). Negative symptom and hypomania groups did not differ significantly (*p* = .111) (Fig 2).

**Fig 2.**
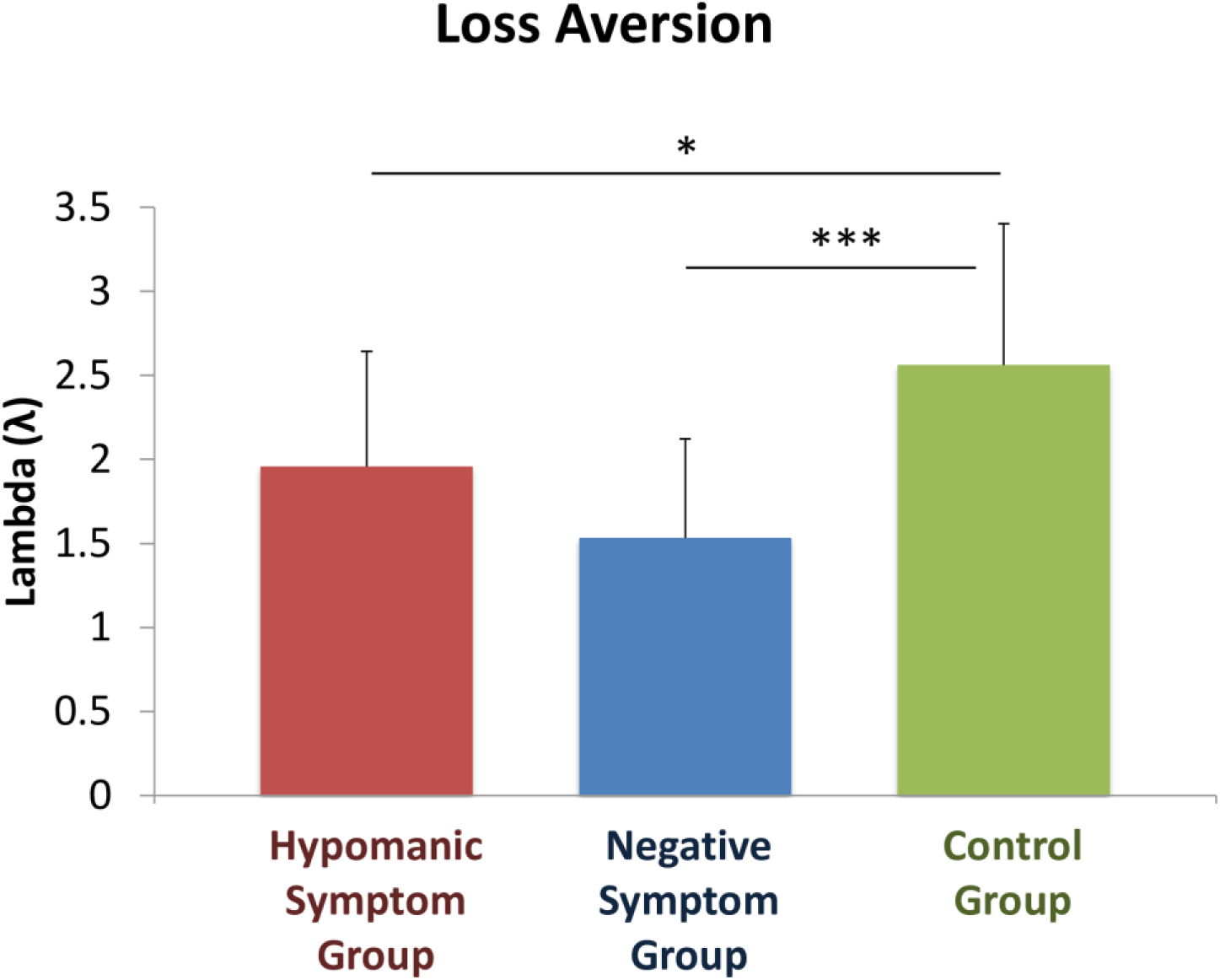
Loss aversion (lambda) in hypomania, negative symptom and control groups. Data were analyzed by a one-way ANOVA (*p* < .001), and paired comparisons were done with LSD test (indicated by \*\*\**p* < .001; \**p <* .05). Means are given with Standard Deviations. *N* = 15 per group.

Despite not being the focus of the current study on loss aversion, similar results were obtained for risk aversion (*F*(2,42) = 5.130, p = .010), where post-hoc testing revealed less risk aversion in participants with higher negative symptoms (*p* = .003) or hypomania (*p* = .040) compared to controls and no difference between symptom groups (*p* = .317).

No significant difference was observed regarding consistency (significant main effect of group (*F*(2,42) = 1.418, p = .254).

Based on the observed group difference in age, we conducted a further analysis where age was included as a covariate. There was a significant main effect of group on loss aversion when introducing the covariate age (*F*(2,41) = 9.5135, *p* < .0001). The introduction of the covariate age reduced the previously observed effect of reduced loss aversion in the hypomania group compared to the control group to trend-level (*p* = .056), while a significant difference between hypomania group and negative symptom group arose (*p* = .03), with higher loss aversion in the hypomania group compared to the negative symptom group.

## Discussion

We applied a stratified approach in a non-clinical population to study how the two symptom dimensions of negative symptoms and hypomania relate to loss aversion. We present the first evidence that loss aversion is similarly diminished in participants with subclinical negative and hypomania symptoms. This suggests that these distinct symptom dimensions manifest in a common behavioral phenotype.

The association of reduced loss and risk aversion with negative symptoms in our study may be important for understanding previous findings of diminished loss aversion in patients with schizophrenia (of which negative symptoms are a core feature). One such study, using a computerized version of the iterated prisoners’ dilemma [21], demonstrated diminished loss aversion in patients with schizophrenia. Furthermore, the extent of loss aversion in patients with schizophrenia was found to be negatively correlated with schizophrenia illness severity. Less ill patients showed a loss aversion more similar to controls and this correlation with loss aversion was with positive symptoms but not with negative symptoms, based on the Positive And Negative Symptom Scale (PANSS) [35]. This finding is not completely in line with our results of a reduced loss aversion in a stratified subclinical negative symptom group. Another study used a task were subjects were asked to evaluate the price of a mug in a buying and in a selling scenario [22]. Patients with schizophrenia showed less loss aversion in comparison to healthy controls, and this reduction correlated with age, duration of illness and hospitalization and poorer cognitive performance, but not with current psychopathology. Yet another study used two gambling tasks, involving the differentiation between losing and keeping [20]. This study found a reduced sensitivity to losses (increased tendency to gamble when faced with a certain loss) in patients with schizophrenia compared to healthy controls. Nevertheless, an even greater reduction in sensitivity to gains was found, which indicates no significant difference in estimated loss aversion between schizophrenia patients and controls [21]. To summarize, reduced sensitivity to losses was demonstrated to be a feature of schizophrenia in the above mentioned studies in line with our findings. Still, it has to be considered that different task paradigms and scales to assess negative symptoms were used, which might complicate the interpretation and comparability of results and also explain the absence of loss aversion in the last study [20]. Furthermore, one has to consider, that in the above mentioned studies, patients with clinical symptoms participated, while the present study investigated a non-clinical sample.

The theory of altered salience, i.e. abnormal weighing of stimuli [21,36], offers one possible explanation for a reduced loss aversion in association with schizophrenic negative symptoms. In other words, the salience of loss is reduced via competitive interference from other external and internal stimuli, which might impede a motivational behavior directed at avoiding losses. This hypothesis is supported by the lack of a framing effect in schizophrenia patients [20], i.e. the context seems not to influence the decision-making in schizophrenia as much as it does in healthy controls.

Depression is a diagnostic category with considerable clinical overlap with negative symptoms, and that’s why one could have expected similar findings in depression and negative symptoms. Nevertheless, several studies observed an increased loss aversion in depression in decision-making tasks [37,38]. The crucial differences in loss aversion found in depression and negative symptoms that we emphasize here, could be explained by different underlying mechanisms of reactivity to loss: Depressed patients show an increased reactivity to distress [39,40], while patients suffering from primary negative symptoms exhibit a reduced sensitivity to negative (distressing) stimuli [41]. Still, this theory includes a highly reduced salience in patients with primary negative symptoms and is therefore pointing to another direction than the above postulated theory of an aberrant salience in negative symptoms.

In the hypomania group, loss aversion was also reduced, which can be interpreted in the context of the diagnostic definition of a manic episode. This definition includes impulsivity and therefore suggests reduced loss aversion [1]. Interestingly, manic patients show a reduced perception of loss as such [42,43], a feature that is comparable to the absence of distress in schizophrenia. This supports the finding of a reduced loss aversion in both negative symptoms and mania in comparison to an increased loss aversion and reactivity to negative stimuli in depression. Nevertheless, the relationship between manic episodes and loss aversion needs further investigation, including also repetitive assessments in different psychopathologic states over time.

Given the opposing nature in terms of activity and hedonia level between negative symptoms and (hypo-) mania [1], one could have expected intuitively different effects on loss aversion in the two groups. Behavioral finance theories investigating portfolio selection strategies suggests that depressed patients aim for a minimal loss, while manic patients aim for a maximal gain, indifferent of incurring losses in their choice behavior [43,44]. Based on this theory, one could have expected an increased loss aversion in negative symptoms (given the high clinical overlap of negative symptoms and depression) and a reduced loss aversion in mania. The contrasting findings in this study of a reduced loss aversion in both groups might be accompanied by separate underlying mechanisms.

Despite not being the focus of the current study, we investigated further also the parameters risk aversion and consistency: Risk aversion was reduced in negative symptom and hypomania groups, similar to loss aversion. These findings can as well be interpreted in the context of an aberrant salience hypothesis for negative symptoms, where the detection of risk is also impaired due to noise, similar to the detection of potential loss. However, group differences in consistency do not reach significance level, probably due to small sample sizes in the current study. Contrastingly, in previous studies, risk aversion was demonstrated to be increased in schizophrenia compared to healthy controls. Furthermore, risk aversion was also demonstrated to be increased in depressed patients, which is in line with an increased loss aversion in depression and suggests that the mechanisms of regulating loss suggesting that the mechanisms underlying risk aversion are regulated differently [45,46].

Our study does have some limitations. We have a small sample size and an absence of measures on ethnicity and did not assess subclinical positive symptoms of participants. Despite these limitations, we believe that the current study presents an important approach to investigating altered decision-making in sub-clinical populations.

Our study demonstrated an inverse relationship of both hypomania and negative symptom dimensions to loss aversion. This suggests that these distinct symptom dimensions manifest a common behavioral phenotype, which is highly relevant to the understanding of altered decision-making. Open research questions to be addressed in future studies include the following: What is the influence of the context or “frame” in which a decision is taken? A recent critique on loss aversion suggested a dependence of loss aversion depending on the context rather than being a general principle [47]. What role does emotion regulation, i.e. the identification and modification of emotions via a cognitive process, which is altered in negative symptoms and (hypo-) mania, play in decision-making processes? Interestingly, an increased emotion regulation was demonstrated to reduce loss aversion [34,48]. And finally the question needs to be addressed, if an altered loss aversion is inherently maladaptive?

## Data Availability

Data are at available at the Harvard Dataverse: https://doi.org/10.7910/DVN/W2KN1M

## Acknowledgements

We thank Nicole Hauser for her help in data acquisition.

